# Durability of antibody responses elicited by a single dose of Ad26.COV2.S and substantial increase following late boosting

**DOI:** 10.1101/2021.08.25.21262569

**Authors:** Jerald Sadoff, Mathieu Le Gars, Vicky Cardenas, Georgi Shukarev, Nathalie Vaissiere, Dirk Heerwegh, Carla Truyers, Anne Marit de Groot, Gert Scheper, Jenny Hendriks, Javier Ruiz-Guiñazú, Frank Struyf, Johan Van Hoof, Macaya Douoguih, Hanneke Schuitemaker

## Abstract

**Background:** We evaluated the durability of SARS-CoV-2 antibody levels elicited by the single dose Janssen COVID-19 vaccine, Ad26.COV2.S, and the impact on antibody responses of boosting with Ad26.COV2.S after 6 months in clinical trial participants.

**Methods:** Spike-binding antibody and SARS-CoV-2 neutralizing antibody levels elicited by a single-dose Ad26.COV2.S (5×10^10^ viral particles [vp]) primary regimen and booster doses (5×10^10^ vp and 1.25×10^10^ vp) were assessed by ELISA and wild-type VNA in sera from participants in a Phase 1/2a clinical trial (Cohort 1a, 18–55 years old, N=25; Cohort 2a, 18–55 years old boosted at 6 months, N=17; Cohort 3, ≥65 years old, N=22) and a Phase 2 clinical trial (18–55 and ≥65-year old participants boosted at 6 months, total N=73). Neutralizing antibody levels were determined approximately 8 months after the primary vaccination in participants aged 18–55 years and approximately 9 months in participants aged ≥65 years. Binding antibody levels were evaluated 6 months after primary vaccination and 7- and 28-days after booster doses in both age groups.

**Results:** A single dose of Ad26.COV2.S elicited neutralizing antibodies that remained largely stable for approximately 8–9 months and binding antibodies that remained stable for at least 6 months irrespective of age group. A 5×10^10^ vp booster dose at 6 months post prime vaccination in 18–55-year-old adults elicited a steep and robust 9-fold increase at Day 7 post boost compared to Day 29 levels following the initial immunization. A lower booster dose of 1.25×10^10^ vp at 6 months in adults 18–55 and ≥65 years of age also elicited a rapid and high increase of 6–7.7 fold at Day 28 post boost compared to Day 29 levels following the initial immunization, with similar magnitude of post-boost responses in both age groups.

**Conclusions:** A single dose of Ad26.COV2.S, which demonstrated protection in a Phase 3 efficacy trial, elicited durable neutralizing and binding antibodies for at least 8 and 6 months, respectively, in adults >18 years of age at levels similar to Day 29 responses. A 5×10^10^ vp or 1.25×10^10^ vp booster dose at 6 months elicited rapid and robust increases in spike binding antibody levels. The anamnestic responses after booster immunization imply robust immune memory elicited by single-dose Ad26.COV2.S.

## Introduction

Severe acute respiratory syndrome coronavirus 2 (SARS-CoV-2) is responsible for the ongoing COVID-19 pandemic. Janssen’s COVID-19 vaccine, Ad26.COV2.S,^1^ has been demonstrated to be safe, immunogenic, and to confer high protective efficacy against severe-critical COVID-19 disease and COVID-19–related hospitalization and death.^2,3^ However, as already observed for one of the other COVID-19 vaccines,^4–6^ vaccine-mediated protection against COVID-19 may decline with time. This decline may imply that antibody levels are waning or are less effective against emerging variants of concern, that immune priming has elicited insufficient immune memory to support anamnestic responses upon exposure to SARS-CoV-2, or a combination of these. Longitudinal evaluation of immune responses following primary vaccination gives insight into the durability of SARS-CoV-2–specific immunity and the potential need for a booster dose to further increase antibody titers at or above protective levels and to enhance immune memory. High antibody (Ab) titers are even more important in the context of emerging variants of concern that are relatively resistant to antibody mediated neutralization^7–9^ and for which antibody levels may need to be higher to confer protection against acquisition and mild-to-moderate disease, especially in populations at high risk for COVID-19, such as the elderly population.

Here, in Phase 1/2a and Phase 2 clinical trial participants 18–55 and ≥65 years of age, we evaluated the durability of SARS-CoV-2 neutralizing antibody and binding antibody levels elicited by a single dose of Ad26.COV2.S at 5×10^10^ vp as a primary vaccination. Moreover, we studied the impact of homologous boosting with Ad26.COV2.S at a 5×10^10^ vp dose 6-months post primary vaccination on these antibody levels. In addition, we tested the impact of a 4-fold lower booster dose of Ad26.COV2.S at 6 months post primary vaccination to assess the robustness of immune memory elicited by the single dose primary regimen, both in younger and older adults. Overall, Ad26.COV2.S-elicited immunity is durable for at least 8 months and can be further increased with a booster in adults >18 years of age.

## Methods

### Study participants and immunogenicity assessment

Participants from an ongoing Phase 1/2a study (COV1001, NCT04436276; Cohort 1a, 18–55 years old [N=25]; Cohort 2a, 18–55 years old [N=17]; Cohort 3, ≥65 years old [N=22]) and an ongoing Phase 2 study (COV2001; NCT04535453; 18–55 and ≥65 years old [total N=73]) received a single dose of Ad26.COV2.S (5×10^10^ viral particles [vp]) as a primary regimen. In addition, Phase 1/2a Cohort 2a and Phase 2 participants received homologous booster doses of Ad26.COV2.S of 5×10^10^ vp or 1.25×10^10^ vp, respectively, at 6 months post primary vaccination. Spike-binding and neutralizing antibody levels were assessed by ELISA during a 6- to 9-month follow up after a single dose Ad26.COV2.S primary regimen, and after a booster dose at 6 months post primary regimen. Neutralizing antibody titers were evaluated in a subset of participants from Phase 1/2a Cohorts 1a and 3 and Phase 2. Binding antibody geometric mean concentrations (GMCs) and neutralizing antibody geometric mean titers (GMTs) were determined per study protocols and amendments at various days based on regimen and age group after primary vaccination, as shown in Figures 1–3. Binding antibody GMCs were uniformly evaluated at days 7 and 28 after booster doses.

**Figure 1:**
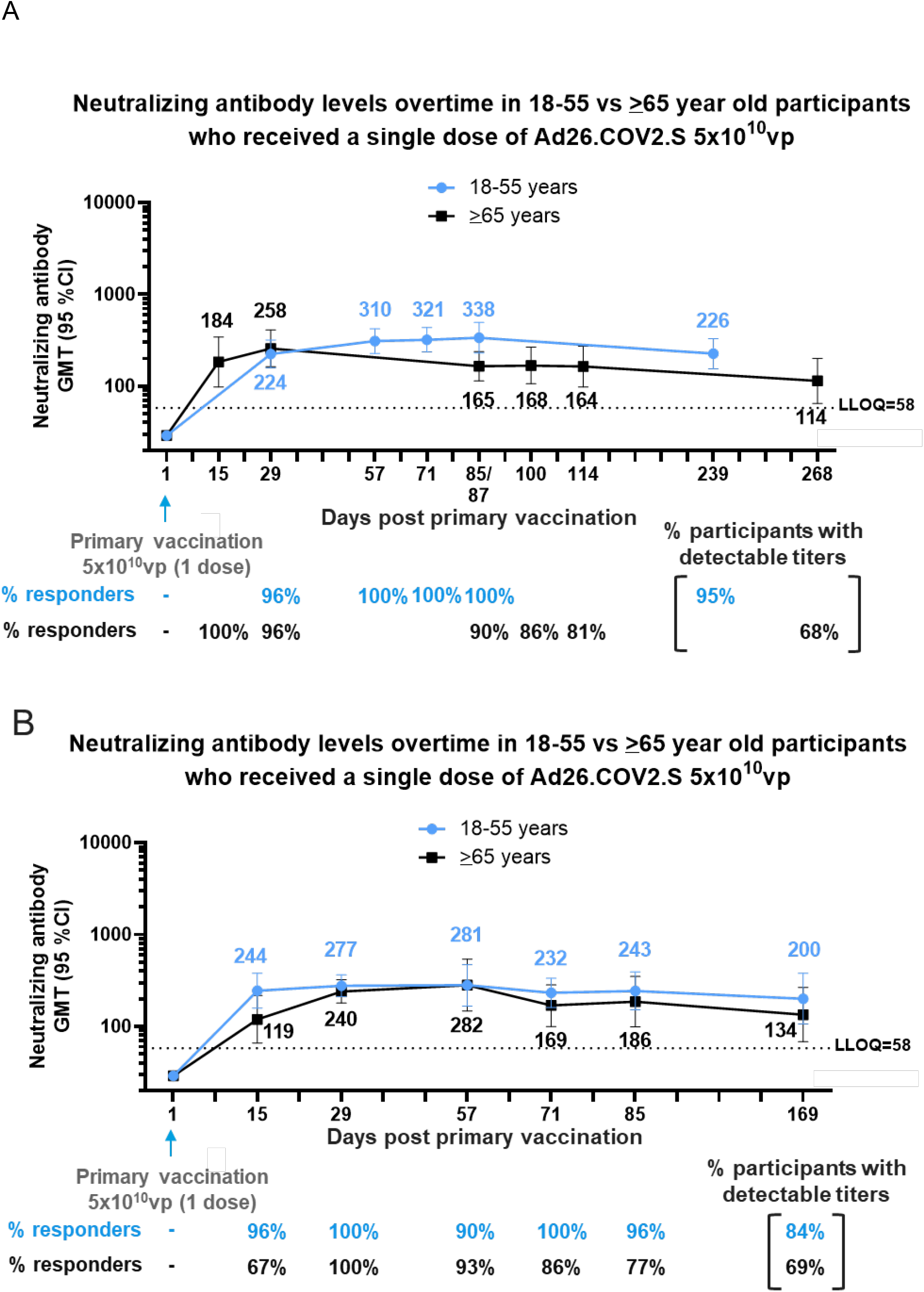
Durability of neutralizing antibody responses following a single dose of Ad26.COV2.S (5×10^10^ vp) in participants 18–55 and ≥65 years old, respectively, from Phase 1/2a and 2 clinical trials. A) Phase 1/2a participants, 18–55 and ≥65 years of age, were administered a single dose of Ad26.COV2.S (5×10^10^vp) at Day 1, as primary vaccination. Serum neutralizing antibody responses against SARS-CoV-2 were evaluated by wtVNA up to 8 to 9 months post primary vaccination, in a subset of 18–55 (N=25) and ≥65 (N=15)-year old participants. B) Phase 2 participants, 18–55 and ≥65 years of age, were administered a single dose of Ad26.COV2.S (5×10^10^ vp) at Day 1, as primary vaccination. Serum neutralizing antibody responses against SARS-CoV-2 were evaluated by wtVNA, up to 6 months post primary vaccination in a subset of 18–55 (N=25) and ≥65 (N=22) year old participants, respectively. Participants 18–55 and ≥65 years of age are represented with blue and black lines, respectively. GMTs are depicted above each time point and response rates are illustrated at the bottom of each panel.

A participant was considered a responder after primary vaccination regimen if: baseline antibody titers were below the lower limit of quantification (LLOQ) prior vaccination and were above the LLOQ after vaccination; or if baseline antibody titers were above LLOQ prior to vaccination and were 4-fold higher than baseline titers after vaccination.

All participants were monitored for solicited and unsolicited local and systemic adverse events (AEs) by 7-day diary cards and 28-day follow-up for unsolicited AEs. Serious adverse events (SAEs) and AEs of special interest were collected throughout each study.

All relevant ethical guidelines have been followed, all necessary IRB and/or ethics committee approvals have been obtained, all necessary patient/participant consent has been obtained and the appropriate institutional forms archived. The COV1001 study was reviewed and approved by local ethics committees (Comité d’Ethique Hospitalo-Facultaire Sain-Luc, Université Catholique de Louvain on July 16, 2020) and institutional review boards (IRB) (approval by Advarra IRB on June 29 and July 10, 2020, for New Orleans Center for Clinical Research and Optimal Research sites, respectively). The COV2001 study was reviewed and approved by local ethics committees (Ethik-Kommission der Ärztekammer Hamburg on August 19, 2020; Landesamt für Gesundheit und Soziales Berlin Geschäftsstelle der Ethik-Kommission des Landes Berlin on August 26, 2020; Ethikkommission an der Medizinischen Fakultät der Universität Rostock on September 01, 2020; and Raad van Bestuur UMC Utrecht on August 27, 2020) and regional ethics committees (Ethikkommission an der Medizinischen Fakultät der Universität Rostock on September 01, 2020 and September 03, 2020; Comité Ético de Investigación Clínica-Regional de la Comunidad de Madrid on August 28, 2020; and Central Committee on Research Involving Human Subjects (CCMO) on August 26, 2020. All participants provided written informed consent before enrollment. The trials adhere to the principles of the Declaration of Helsinki and to the Good Clinical Practice guidelines of the International Council for Harmonisation.

### SARS-CoV-2 wild-type virus neutralization assay

Neutralizing antibodies capable of inhibiting wild-type virus infections were quantified using the wild-type virus microneutralization assay (MNA) that was developed and qualified by Public Health England (PHE). The virus stocks used were derived from the Victoria/1/2020 strain, and the LLOQ of the assay is 58 IC_50_.

In brief, 6 two-fold serial dilutions of heat-inactivated human serum samples were prepared in 96-well transfer plate(s). The SARS-CoV-2 wild-type virus was subsequently added to the serum dilutions at a target working concentration (approximately 100 plaque-forming units [PFU]/well) and incubated at 37°C for 60 to 90 minutes. The serum-virus mixture was then transferred onto assay plates previously seeded overnight with Vero E6 African green monkey kidney cells and incubated at 37°C and 5% CO_2_ for 60 to 90 minutes before the addition of carboxymethyl cellulose (CMC) overlay medium and further incubation for 24 hours. Cells were then fixed and stained using an antibody pair specific for the SARS-CoV-2 RBD spike (S) protein, and immunoplaques were visualized using TrueBlue™ substrate (KPL, Milford, MA, USA).

Immunoplaques were counted using the Immunospot Analyzer (Cellular Technology Limited, Cleveland, OH, USA). The immunoplaque counts were exported to SoftMax Pro (Molecular Devices, San Jose, CA, USA) and the neutralizing titer of a serum sample was calculated as the reciprocal serum dilution corresponding to the 50% neutralization antibody titer (IC_50_) for that sample.

### Spike protein enzyme-linked immunosorbent assay (ELISA)

SARS-CoV-2 pre-spike-specific binding antibody concentrations were determined using the human SARS-CoV-2 pre-spike IgG ELISA, an indirect ELISA that is based on antibody/antigen interactions. The SARS-CoV-2 antigen was a stabilized pre-fusion spike protein ([2P], Δfurin, T4 foldon, His-tag), derived from the first clinical isolate of the Wuhan strain (Wuhan, 2019, whole genome sequence NC_045512), produced in ES-293 cells. The ELISA was developed and qualified for human serum at Nexelis, (Laval, PQ, Canada). The lower limit of quantification of the assay is 53 ELISA Units (EU/mL).

In brief, purified SARS-CoV-2 pre-spike antigen was adsorbed to the wells of a microplate and diluted serum samples (test samples, standard, and quality controls) were added. Unbound sample was washed away, and enzyme-conjugated anti-human IgG added. After washing excess conjugate away, 3,3′,5,5′-tetramethylbenzidine (TMB) colorimetric substrate was added. After the established time period, the reaction was stopped. A reference standard on each test plate was used to quantify the amount of antibodies against SARS-CoV-2 pre-spike in the sample according to the unit assigned by the standard (EU/mL).

## Results

### Durability of humoral immunity after single dose Ad26.COV2.S (5×10^10^ vp)

We previously reported the short-term immunogenicity after single-dose primary vaccination with Ad26.COV2.S in participants of our Phase 1/2a study.^2^ Here we report the levels of neutralizing antibodies after longer follow up of the participants of Cohort 1a (18–55 years old; 8 months follow-up) and Cohort 3 (≥65 years old; 9 months follow-up). In participants aged 18–55 years old (Cohort 1a), neutralizing antibody responses were detectable up to at least Day 239 (8 months) after a single dose of Ad26.COV2.S, with GMTs of 226 and 21/22 (95%) of participants with detectable titers, which is similar to what was observed by Day 29 post dose 1 (GMT of 224 and 96% of participants with positive titers; Figure 1A). In participants ≥65 years of age (Cohort 3), neutralizing antibodies were still detectable in 13/19 (68%) of participants by Day 268 (9 months) after one dose of Ad26.COV2.S, with GMTs of 114. This represents a 2.3-fold decrease in GMTs compared to Day 29 post dose 1 (GMT of 258) in participants ≥65 years of age (Figure 1A).

From Cohort 2a of our Phase 1/2a study, which is described in the supplementary material of Sadoff et al., 2021,^2^ 17 participants 18–55 years of age had undetectable binding antibody levels at Day 8 post prime vaccination but could be measured at the next timepoint (Day 29; GMC, 418 [95% CI, 322–554], with 100% responders; Figure 2A and 2B). This is in the same range as the observation in adults 18–55 years old in our Phase 2 study (Figure 3). By month 6, GMCs in participants in Cohort 2a had increased to 900 (441–1643) with 100% of participants still having detectable binding antibodies (Figure 2A).

**Figure 2:**
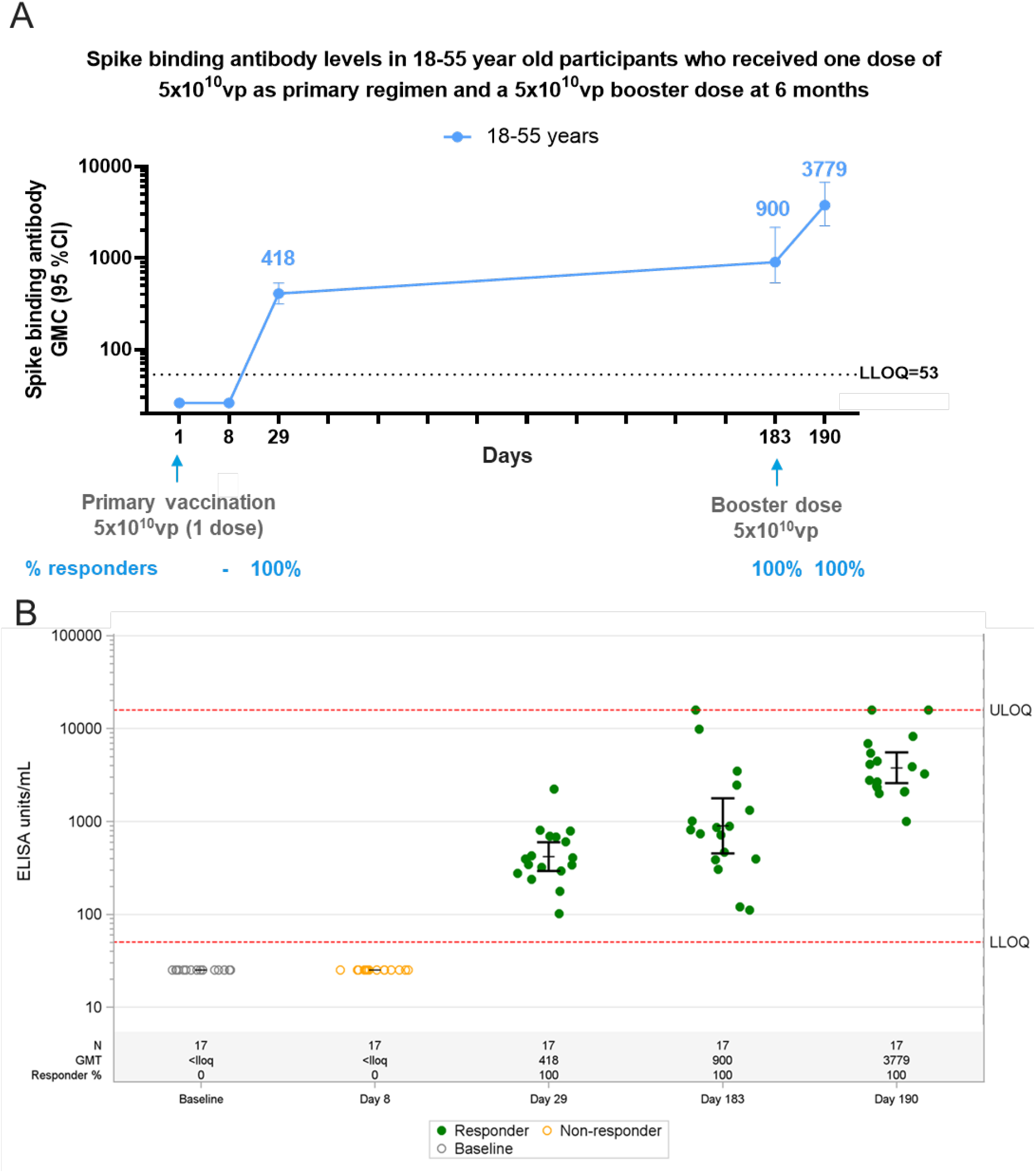
Durability of spike binding antibody responses up to 6 months following a single dose of Ad26.COV2.S (5×10^10^ vp) in and impact of a booster dose at 6 months post primary vaccination, in18–55 year-old participants from a Phase 1 clinical trial. Phase 1 participants, age 18–55 (N=27) years of age, were administered a single dose of Ad26.COV2.S (5×10^10^ vp) at Day 1 and 20 participants received a booster dose of Ad26.COV2.S (5×10^10^ vp) at approximately 6 months (Day 183) post primary vaccination and 17 participants had data available at Day 190. Serum spike binding antibodies against SARS-CoV-2 were evaluated in an S-ELISA up to 7 days post boost (Day 190). A) Participants18–55 years of age are represented with a blue line. GMCs are depicted above each time point and response rates are illustrated at the bottom of each panel. B) Dot plots representing the distribution of the participants per timepoint. N, GMC (ELISA Unit/mL) and percent responders are represented at the bottom of each plot. Grey open circles represent baseline binding antibody levels, green dots indicate that participants are responders and yellow open circles indicate that participants are non-responders.

**Figure 3:**
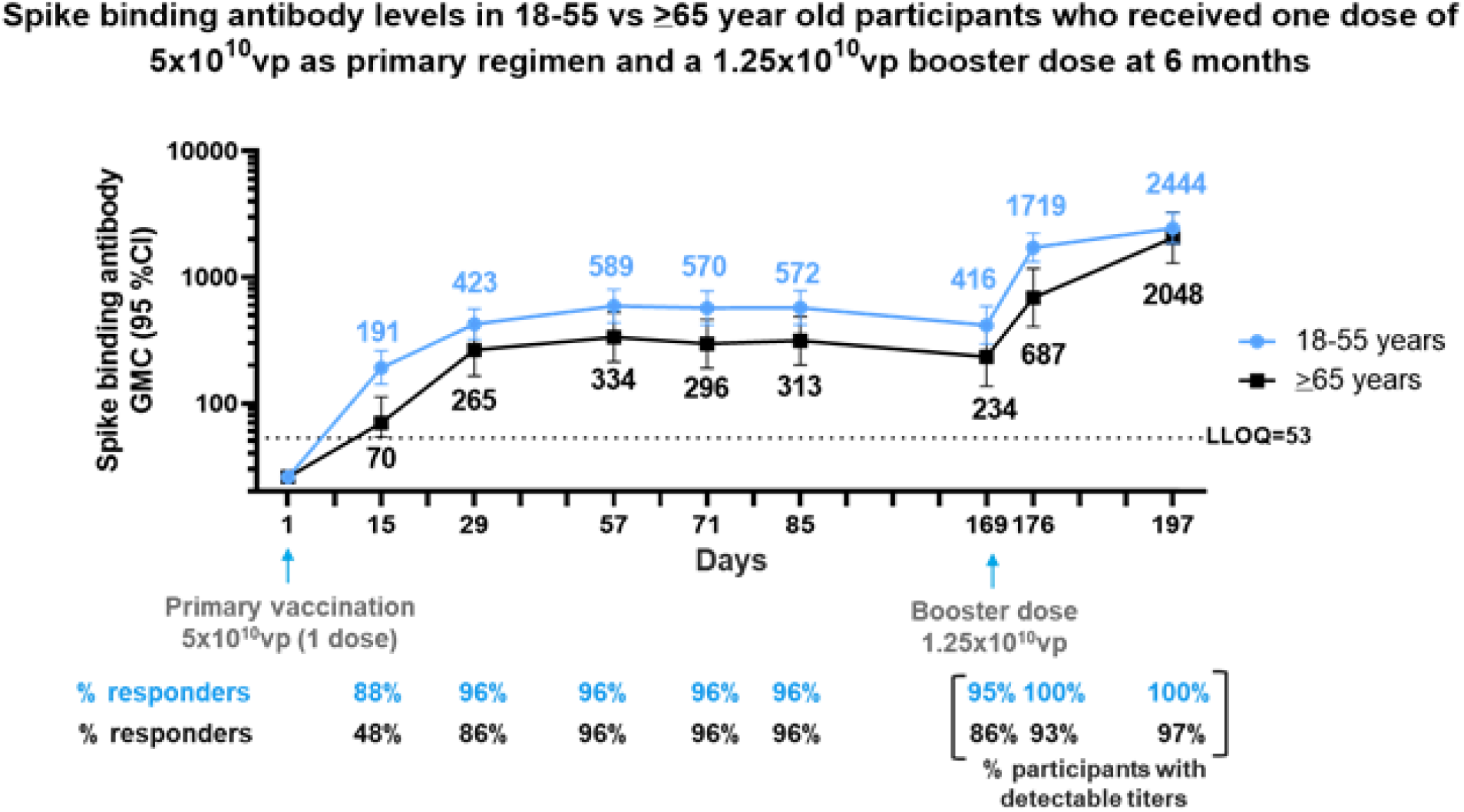
Durability of spike binding antibody responses 6 months after primary immunization with a single dose of Ad26.COV2.S (5×10^10^ vp) and booster responses 7 and 28 days later in 18–55 and ≥65-year-old participants from a Phase 2 clinical trial. Phase 2 participants 18–55 (N=50) and ≥65 (N=25) years of age, were administered a single dose of Ad26.COV2.S (5×10^10^ vp) as primary vaccination at Day 1 and 73 received a booster dose of Ad26.COV2.S (1.25×10^10^ vp) at 6 months (Day 169) post primary vaccination. Serum spike binding antibody responses against SARS-CoV-2 were evaluated by S-ELISA up to 28 days post boost (Day 197). Participants 18–55 and ≥65 years of age are represented with blue and black lines, respectively. GMCs are depicted above each time point and response rates are illustrated at the bottom of each panel.

In a Phase 2 study (COV2001), a single dose of Ad26.COV2.S elicited neutralizing antibody responses by Day 15 in 21 of 22 participants aged 18–55 years (96% responders; GMT, 244 [158–277]) and in 10 of 15 participants ≥65 years of age (67% responders; GMT, 119 [66–217]; Figure 1B). These responses further increased by Day 29 in both age groups (18–55 years, 100% responders and GMT, 277 [211–365]; ≥65 years, 100% responders and GMT, 240 [179–322]; Figure 1B). Up to Day 85, neutralizing antibody responses remained stable in participants aged 18–55 years (96% responders and GMT, 243) while they decreased modestly to a GMT of 186 (99–349) in participants ≥65 years old, with 77% still above the LLOQ of the assay. Neutralizing antibody levels in participants 18–55 years old 6 months after vaccination were in a similar range as Day 29 levels (GMT, 200 [106–378]; 84% responders). In adults ≥65 years old, the GMT of neutralizing antibody at 6 months after primary vaccination was 134 (68–266) with 69% responders.

Binding antibody levels also gradually increased from baseline to Day 15 to Day 29 and remained stable up to Day 85 in both age groups (18–55 years: GMC, 572 [420–780], ≥65-years: GMC, 313 [201–486], with 96% above LLOQ of the assay in both groups; Figure 3). GMCs in participants ≥65 years old were slightly lower at all timepoints compared to those aged 18–55 years. At 6 months after primary vaccination, GMCs of binding antibodies had declined to 416 (294–588) and 234 (136–403) with 93% and 86% of participants still having titers above LLOQ of the assay in those aged 18–55 and ≥65 years, respectively. Binding antibodies at 6 months post primary vaccination were undetectable in 2 of 44 (5%) participants aged 18–55 years and in 4 of 29 (14%) participants ≥65 years old.

### Humoral immune responses after homologous boosting with Ad26.COV2.S at a 5×10^10^ vp dose level

Clinical trial participants in our Phase 1/2a study (18–55 years of age; N=17) who had received a primary vaccination with a single dose of Ad26.COV2.S at a dose level of 5×10^10^ vp were given a homologous booster vaccination 6 months later with a dose of 5×10^10^ vp. By day 7 post boost, all participants demonstrated a robust increase in binding antibody levels of 4.7-fold (GMC, 3779 [2741–4243]) compared to immediate pre-boost binding antibody levels and a 9-fold increase in GMC compared to Day 29 binding antibody levels after the initial immunization (Figure 2).

### Humoral immune responses after low-dose homologous boosting with Ad26.COV2.S

In the Phase 2 trial, a lower homologous booster dose of 1.25×10^10^ vp Ad26.COV2.S was given at 6 months in 44 participants 18–55 years of age and 29 participants ≥65 years of age (Figure 3). This lower booster dose also elicited a rapid and high increase of 3.6-fold increase in binding antibody levels at 7 days post boost (GMC, 1719 [1321–2236]) as compared to immediate pre-boost binding antibody levels. Levels of antibodies further increased by Day 28 post boost (GMC, 2444 [1855–3219]), representing 6.4-fold and 6.9-fold increases compared to Day 29 levels after the initial immunization and immediate pre-boost antibody levels, respectively. While the kinetics of the post low-dose booster were slower in the ≥65-year-old adults, the magnitude of the response by Day 28 was similar in both younger and older adults studied (Figure 3).

### Safety of a booster dose of Ad26.COV2.S

In Cohort 2 of the Phase 1/2a study (COV1001) in 17 participants, both post-primary regimen and post-booster reactogenicity appeared similar as compared to the previously reported Cohort 1 reactogenicity.^2^ Reactogenicity following the boost may have been more comparable to the reactogenicity previously reported from Cohort 1 following a second dose at 57 days after the initial dose, which was lower than following the initial priming dose, but the limited number of participants precludes precision in these measurements.

In the Phase 2 study (COV2001), when booster doses of 1.25×10^10^ vp Ad26.COV2.S were given at 6 months after the primary single-dose immunization of 5×10^10^ vp in 81 participants, solicited AEs after the primary dose versus after the booster were 67.9% versus 54%, respectively, and for grade 3 solicited AEs, 1.2% versus 0%, for solicited local AEs, 51.9% versus 47%, for solicited local AEs of grade 3 or more 0% versus 0%, for solicited systemic AEs, 66.7% versus 28%, and for solicited systemic AEs of grade 3 or more, 1.2% vs 0%.

## Discussion

We recently reported that a single dose of the Janssen Ad26.COV2.S COVID-19 vaccine is immunogenic and highly efficacious against severe COVID-19 disease and COVID-19 related hospitalization and death, also in areas where the beta variant of concern had high prevalence.^2,3^ In addition, data from the Sisonke study in South Africa among 480,000 health care workers,^10^ which was performed in the context of the current surge with the Delta variant, demonstrates high efficacy of single-dose Ad26.COV2.S against COVID-19 related hospitalization and death due to the Delta variant.

Here, in participants of our Phase 1/2a study, we observed that following a single dose of Ad26.COV2.S (5×10^10^ vp), neutralizing and binding antibody levels remained at or above post-prime levels in the majority of participants, irrespective of age. In the younger age group, antibody levels at 6 months were similar to levels on Day 29 while in older adults the antibody levels showed a slight and insignificant decline between Day 29 and month 6 post vaccination. These durable antibody levels are consistent with our previous observation in a sub-cohort of our Phase 1/2a study where we also demonstrated stable humoral immune responses for 8 months after primary vaccination with Ad26.COV2.S in 18–55 year-old adults, including against Beta and Delta variants of concern.^7^ Together, these data imply that a single dose of Ad26.COV2.S at a dose level of 5×10^10^ vp elicits a high quality and durable immune response of at least 6 to 8 months in adults >18 years of age.

A mechanistic immune correlate of protection has not yet been defined but antibody levels do seem to have some correlation with vaccine efficacy.^11–13^ In our ENSEMBLE Phase 3 efficacy trial, high vaccine efficacy against severe/critical COVID-19 disease was observed as early as Day 7 after vaccination, and by Day 14 after vaccination for prevention of hospitalization and death. GMT of antibodies at 6 to 8 months following immunization were similar, or slightly lower in participants ≥65 years old, to GMTs at 28 days after immunization at a time when protection was seen in our efficacy study, which is similar to results reported earlier in a sub-cohort or our Phase 1/2a study.^7^ It is therefore expected that these durable immune responses may translate into durable vaccine efficacy.

Waning vaccine elicited immune responses and the emergence of virus variants may impact vaccine efficacy.^8,9^ While immune responses were preserved in the majority of participants, we did observe that few participants in both age groups had undetectable levels of SARS-CoV-2 specific neutralizing antibodies at 6 months post primary vaccination with Ad26.COV2.S. Similar or even higher numbers of participants with declining neutralizing antibodies to undetectable levels have been reported for recipients of mRNA based COVID-19 vaccines.^14,15^

We explored here the potential utility of a booster immunization following initial vaccination with Ad26.COV2.S and show that a homologous booster with Ad26.COV2.S (5×10^10^ vp) at 6 months after primary vaccination leads to a rapid increase in spike specific antibody levels. In 18–55 year-old adults, booster doses of either 5×10^10^ vp or 1.25×10^10^ vp gave rapid and strong increases in antibody levels that were significantly higher than antibody levels at Day 29 after primary vaccination, the period in which in our phase 3 efficacy study protection from hospitalization and death by our vaccine was demonstrated, with protection against severe disease commencing already at day 7, at lower levels of antibody than seen at Day 28.^16^ In addition, it is important to emphasize that even older adults in whom spike specific antibody titers had declined to undetectable levels at month 6 post primary vaccination showed similar strong responses to this low dose booster vaccination. These anamnestic responses even in individuals with undetectable antibodies may also occur in response to SARS-CoV-2 infection and hence these people are likely still protected against severe COVID-19.

At this point, only ELISA data were available from samples obtained post booster vaccination and VNA data will become available soon. However, from our previous analyses we know that Ad26.COV2.S-elicited spike specific binding levels strongly correlate with levels of SARS-COV-2 neutralizing antibodies.^2^

These data also confirm our earlier observations^2,17,18^ that homologous boosting with an Ad26-based vaccine works very well, regardless of the induction of antibodies against the vector. Ad26 antibody titers after vaccination are relatively low and can apparently be overcome by even a low dose level of a booster vaccination. It was recently reported that Ad26.COV2.S is also capable of boosting immune responses in people who had received a mRNA based COVID-19 vaccine for primary vaccination.^19^ This data set suggests that Ad26.COV2.S may be utilized to boost both mRNA and Ad26.COV2.S vaccinated individuals. Additional clinical studies are ongoing to confirm the potential of both homologous and heterologous booster strategies with Ad26.COV2.S.

The 9-fold increase in binding antibody levels, observed at Day 7 after boosting at 6 months with the 5×10^10^ vp dose level was greater than the 2.2-fold increase at Day 15 after a second dose at 2 months after the first vaccination.^2^ The benefit of a longer interval between vaccine doses on the magnitude of immune responses has been reported previously^20^ and implies that a booster dose at 6–8 months or perhaps even later timepoints may be optimal.

In the previously reported Phase 1/2a study,^2^ solicited and unsolicited local and systemic adverse events were found to be transient and generally mild following a 5×10^10^ vp dose given 2 months after the single priming dose with less severity compared to the initial dose. Boosting at 6 months with either 5×10^10^ vp dose or the 1.25×10^10^ vp dose in a similar way were found to be safe and well tolerated with primarily transient mild systemic and local adverse events. In the current study, while the numbers of participants given the 5×10^10^ vp booster (N=17) was too small to precisely measure the difference between local and systemic AEs following primary immunization versus booster, the number boosted with the 1.25×10^10^ vp dose (N=81) provided the opportunity to compare this difference in a more robust manner. Solicited local and systemic AEs of grade 3 or more were extremely low following primary or booster immunization, with none seen following the booster. Solicited local AEs were similar after the primary vaccination versus the booster (51% vs 47%) despite the difference in dose, while solicited systemic AEs were demonstrably lower after the boost (61.7% vs 28%). This difference may be attributed to the reduction in reactogenicity seen with Ad26.COV2.S as noted above following a second dose and possibly due to the lower dose, although the lower dose had no effect on local reactogenicity. While our current study is much too small to estimate the risk of a booster dose for the very rare thrombosis with thrombocytopenia syndrome (TTS),^21,22^ data obtained with the AstraZeneca vaccine demonstrate that the risk for TTS after a second dose of ChAdOx1 nCoV-19 vaccine is even significantly lower,^23^ an observation that can likely be extended to Ad26.COV2.S.

Overall, our data demonstrate that a single dose of Ad26.COV2.S elicits durable protective immunity for at least 8 months as well as immune memory that allows for robust anamnestic responses following booster vaccination. Boosting leads to rapid increases of humoral immune responses to 9 times the levels achieved on Day 29 following the primary vaccination. Such markedly boosted immune responses may translate into sustained protective efficacy, including against variants of concern.

## Data Availability

The data sharing policy of Janssen Pharmaceutical Companies of Johnson & Johnson is available at https://www.janssen.com/clinical-trials/transparency. As noted on this site, requests for access to the study data can be submitted through Yale Open Data Access (YODA) Project site at http://yoda.yale.edu.

http://yoda.yale.edu

## Acknowledgement/Funding Statement

Supported by Janssen Vaccines & Prevention B.V. in collaboration with the Biomedical Advanced Research and Development Authority, the National Institutes of Health, the Department of Defense, and the COVID-19 Prevention Network. This project has been funded in whole or in part with Federal funds from the Office of the Assistant Secretary for Preparedness and Response, Biomedical Advanced Research and Development Authority, under Contract No. HHSO100201700018C.

